# “In Situ Resistance Insulin – Localized Type 2 Diabetes Mellitus or Type 6 Diabetes Mellitus?”: A Scoping Review

**DOI:** 10.1101/2024.11.02.24316656

**Authors:** Luís Jesuino de Oliveira Andrade, Gabriela Correia Matos de Oliveira, João Cláudio Nunes Carneiro Andrade, Alcina Maria Vinhaes Bittencourt, Luís Matos de Oliveira

**Affiliations:** Departamento de Saúde Universidade Estadual de Santa Cruz, Ilhéus, Bahia, Brazil; Programa Saúde da Família, Bahia, Brazil; Faculdade de Medicina Universidade Federal da Bahia, Salvador, Bahia, Brazil; Escola Bahiana de Medicina e Saúde Pública, Salvador, Bahia, Brazil

**Keywords:** Insulin resistance, Organ-specific insulin resistance, Diabetes mellitus, Scoping review

## Abstract

In the context of type 2 diabetes mellitus (T2DM), the concept of organ-specific insulin resistance (IR) as a localized manifestation has garnered increasing attention. A scoping review was conducted to investigate the clinical relevance of IR confined to individual organs without systemic metabolic implications. Utilizing a methodological framework adapted from Arksey and O’Malley, a comprehensive search of PubMed was performed, focusing on the period between January 1990 and October 2024. The search strategy combined Medical Subject Headings terms and keywords related to IR and specific organs. Notably, while “insulin resistance” yielded a substantial number of results, the subset of “organ-specific insulin resistance” returned a more limited dataset, highlighting a gap in current literature. The systematic selection process encompassed identification, screening, eligibility, and inclusion stages to ensure robust inclusion criteria. This scoping review underscores the importance of exploring organ-specific IR in the diabetic milieu and sets the stage for further research to elucidate its role in the pathogenesis of T2DM. Conclusion: The findings suggest that investigating organ-specific IR in the context of T2DM is a promising avenue for future research to deepen our understanding of disease pathophysiology. Thus, this scoping review answers the following question “In Situ Resistance Insulin - Localized Type 2 Diabetes Mellitus or Type 6 Diabetes Mellitus?”, emphasizing the need for targeted investigations into localized manifestations of IR and their implications for DM management strategies.

## INTRODUCTION

The understanding of tissue-specific insulin resistance (IR) has evolved over the years, with recent advances focusing on the intricate mechanisms underlying this condition.^1^ Our concept of type 6 diabetes mellitus (T6DM) refers to the localized development of IR in specific organs, such as the brain, hypothalamus, pituitary gland, thyroid, lung, heart, liver, pancreas, kidneys, spleen, small intestine, large intestine, muscle, adipose tissue, vessels, ovaries, and testicles, independent of systemic insulin sensitivity.^2^

Recent studies have highlighted the role of pro-inflammatory cytokines, adipokines, and lipotoxicity in promoting IR at the cellular level, contributing to the pathogenesis of T6DM.^3^ Furthermore, evidence suggests that genetic predisposition and epigenetic modifications may also play a significant role in the development of organ-specific IR, influencing the onset and progression of metabolic disorders.^4^

The identification of novel therapeutic targets aimed at mitigating IR in isolated organs represents a promising approach for the treatment of this condition. Targeted interventions focused on modulating specific molecular pathways involved in tissue-specific insulin signaling hold great potential for improving metabolic health and reducing the risk of complications associated with IR.^5^ Additionally, personalized medicine approaches that consider individual variations in genetic and environmental factors may enhance the efficacy of treatments targeting organ-specific IR in patients with metabolic disorders.^6^ Thus, the concept of T6DM emphasizes the importance of exploring tissue-specific mechanisms of IR to develop targeted therapies for improving metabolic health and reducing its related complications.

A scoping review offers a comprehensive exploration of existing research on a particular topic, identifying key concepts, research gaps, and the extent of available evidence. Unlike systematic reviews, scoping reviews are more flexible in their methodology, accommodating a broader range of study designs. By providing a detailed overview of the literature, scoping reviews inform research agendas, policy development, and practice. For instance, a scoping review by Arksey and O’Malley^7^ established a foundational framework for conducting these studies, outlining key steps such as identifying the research question, selecting studies, charting the data, and collating, summarizing, and reporting the results.

Our objective is to characterize T6DM as an organ- and system-specific IR through a scoping review, encompassing the central nervous system, endocrine glands (hypothalamus, pituitary, thyroid, and reproductive system), respiratory system, cardiovascular system, hepatobiliary system, gastrointestinal tract, hematopoietic system, musculoskeletal system, adipose tissue, and vasculature.

## MATERIALS AND METHODS

### Registration

To guarantee transparency and uphold methodological principles, review protocol was written prior to undertaking the review in the OSF Registries under the unique identifier https://doi.org/10.17605/OSF.IO/WRT8V. This publicly accessible record, available at https://archive.org/details/osf-registrations-wrt8v-v1 comprehensively details the search strategy.

### Search Strategy

A scoping review was performed to explore the clinical significance of organ-specific IR. Following the methodological approach detailed by Arksey and O’Malley,^7^ a comprehensive search of the PubMed literature was conducted to address the question: Can IR confined to individual organs, without systemic metabolic complications, be considered an in situ manifestation of the diabetic state?

This scoping review adhered to the adapted Preferred Reporting Items for Systematic Reviews and Meta-Analyses (PRISMA) checklist and its extension for scoping reviews (PRISMA-ScR),^8^ to ascertain that the review encompassed all pertinent subject areas. The literature search encompassed the PubMed databases, focusing on articles published between January 1990 and October 2024. The search strategy included a combination of Medical Subject Headings (MeSH) terms and keywords, such as “insulin resistance”, “organ-specific insulin resistance”, and “brain”, “hypothalamus”, “pituitary”, “thyroid”, “lung”, “heart”, “liver”, “pancreas”, “kidney”, “spleen”, “small intestine”, “large intestine”, “muscle”, “adipose tissue”, “vessels”, “ovary”, “testicle”. Data were extracted using a standardized data extraction form (Figure 1).

**Figure 1.**
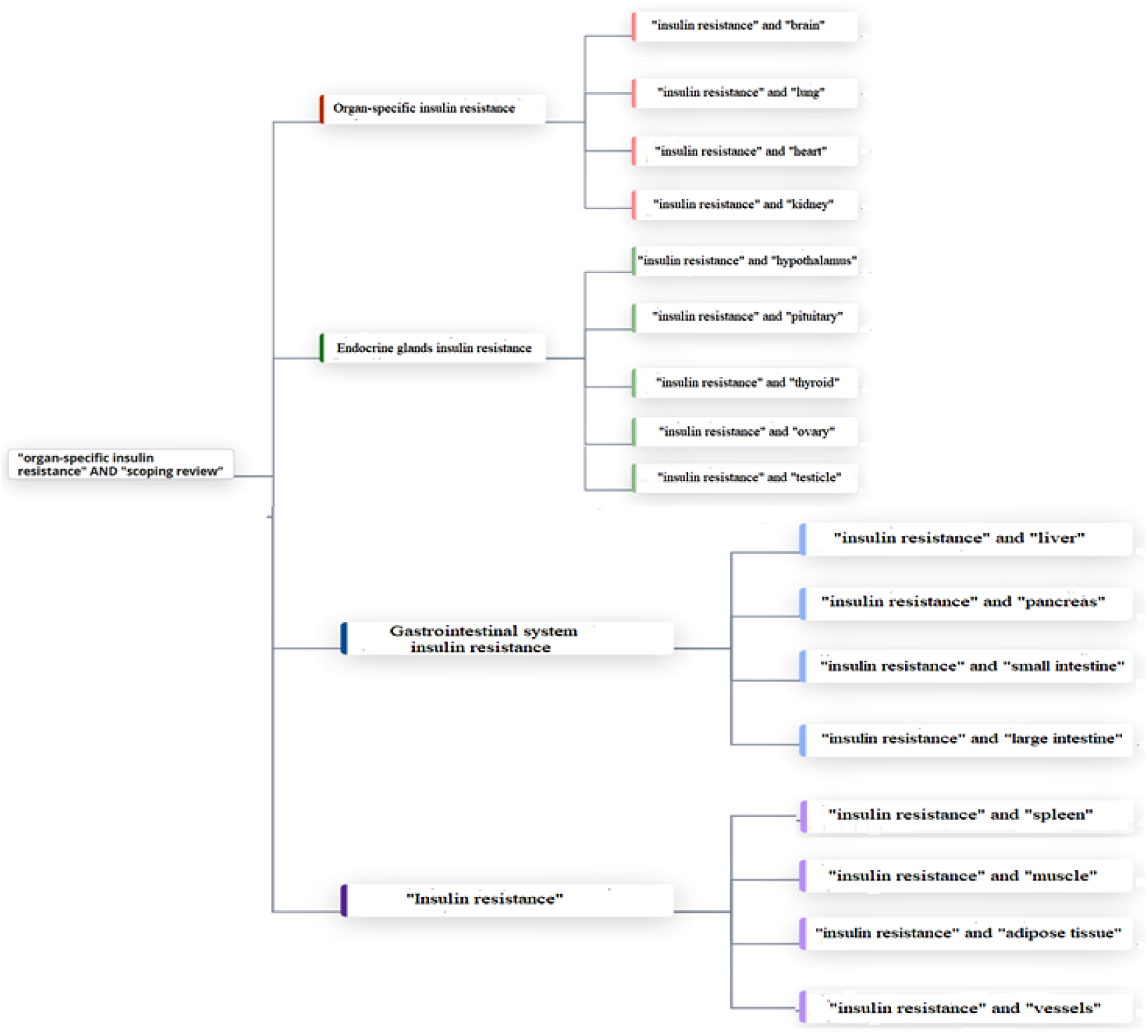
Search strategy

### Inclusion and Exclusion Criteria

All articles within the research domain were meticulously examined. English-language studies explicitly addressing IR were included. Original research articles were considered if they reported IR at the organ or system level. Articles were excluded if they were not human-based studies (e.g., animal research) or if they were published prior to 1990. Review papers, editorials, case studies, and opinion pieces were not included in this analysis.

## RESULTS

A comprehensive literature search was conducted in PubMed using the keywords “insulin resistance” and “organ-specific insulin resistance.” While “insulin resistance” yielded a vast number of results (123,764), “organ-specific insulin resistance” produced a significantly smaller dataset (8 results).

To explore specific organ-system associations, we further refined our search by combining “insulin resistance” with terms representing various organs and systems. When we put the association of the terms “insulin resistance” and “brain” we found 4,504 results, “insulin resistance” and “hypothalamus” we found 1,268 results, “insulin resistance” and “pituitary” we found 1,349 results, “insulin resistance” and “thyroid” we found 1,494 results, “insulin resistance” and “lung” we found 1,185 results, “insulin resistance” and “heart” we found 10,571 results, “insulin resistance” and “liver” we found 25,178 results, “insulin resistance” and “pancreas” we found 2,782 results, “insulin resistance” and “kidney” we found 5,457 results, “insulin resistance” and “spleen” we found 296 results, “insulin resistance” and “small intestine” we found 301 results, “insulin resistance” and “large intestine” we found 24 results, “insulin resistance” and “muscle” we found 15,453 results, “insulin resistance” and “adipose tissue” we found 119,272 results, “insulin resistance” and “vessels” we found 917 results, “insulin resistance” and “ovary” we found 6,614 results, and “insulin resistance” and “testicle” we found 9 results only. We selected only articles that presented a significant association between the terms used.

The selection of articles for this review followed a four-stage process: Identification, Screening, Eligibility, and Inclusion (depicted in Figure 2).

**Figure 2.**
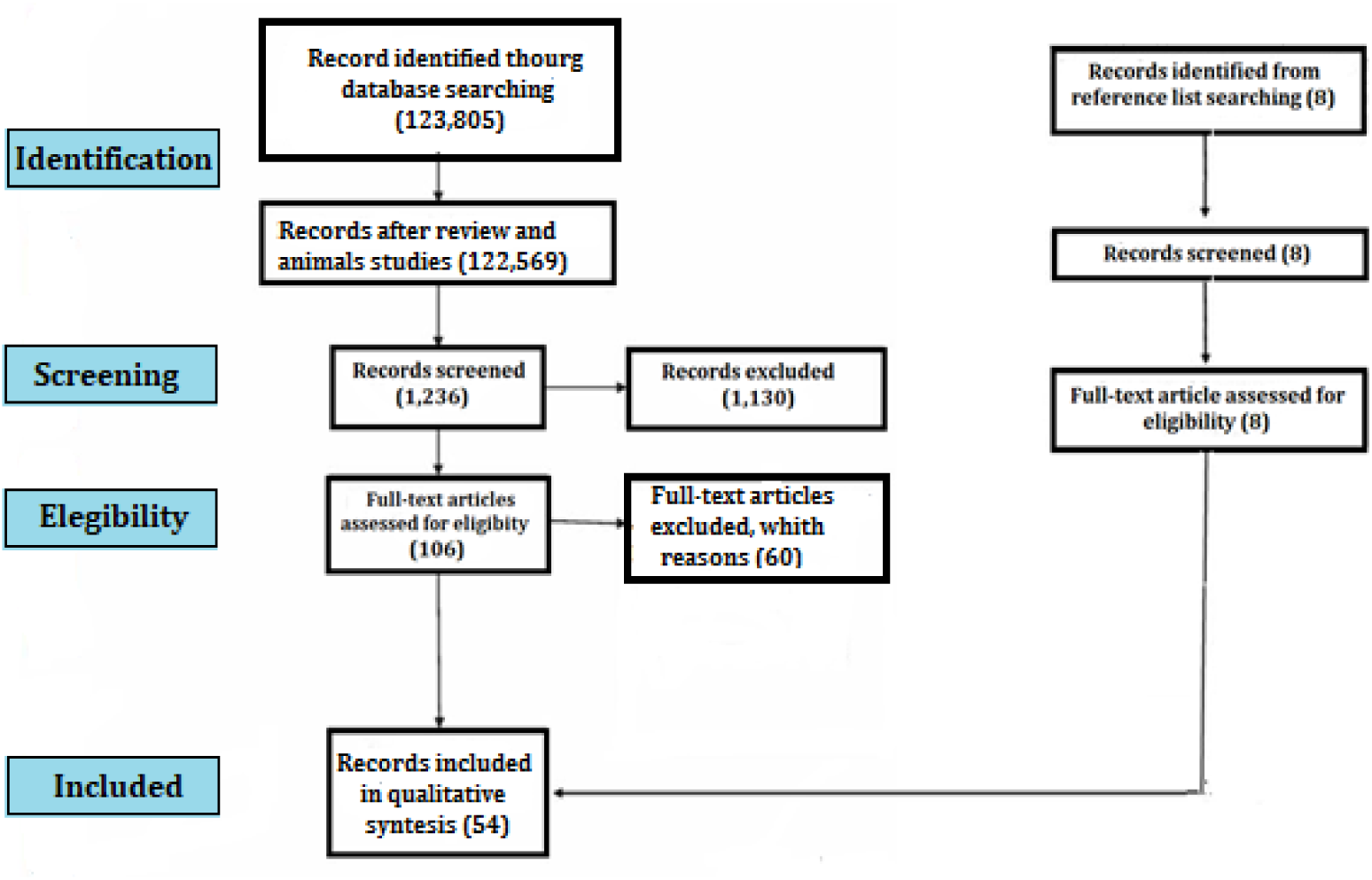
The Process of Selection.

During the identification phase, we identified a pool of potential records from relevant databases and excluded those that did not meet the initial criteria. During the screening stage, we reviewed each record in detail, excluding those deemed ineligible based on our inclusion and exclusion criteria. We then sought to retrieve the full texts of the remaining records, but were unable to obtain some for various reasons. Finally, we assessed the retrieved reports for eligibility and excluded those that did not meet the inclusion criteria. The inclusion stage represented the final set of studies included in the scoping review.

- ***Brain insulin resistance***
  1. Kullmann S, et al. (2022)^9^: Objective: “To evaluate the effects of an exercise intervention on insulin sensitivity of the brain”; Study type: Clinical trial; Conclusion: “An 8-week exercise intervention in sedentary individuals can restore insulin action in the brain”.
  2. Mansur RB, et al. (2021)^10^: Objective: “To directly explore the potential role of neuronal insulin signaling using an innovative technique based on biomarkers derived from plasma extracellular vesicles enriched for neuronal origin”; Study type: Randomized, double-blind, placebo-controlled; Conclusion: “Brain insulin signaling is a target for further mechanistic and therapeutic investigations”.
  3. Nijssen KM, et al. (2024)^11^: Objective: “To investigate longer-term effects of mixed nuts on brain insulin sensitivity in older individuals with overweight/obesity” Study type: Randomized, single-blinded, controlled, crossover trial; Conclusion: “Longer-term mixed nut consumption affected insulin action in brain regions involved in the modulation of metabolic and cognitive processes in older adults with overweight/obesity”.
- ***Hypothalamic insulin resistance***
  4. Kullmann S, et al. (2022)^12^: Objective: “SGLT2 inhibition may be a pharmacological approach to reverse brain insulin resistance?”; Study type: Randomized, double-blind, placebo-controlled clinical trial; Conclusion: “The results corroborate insulin resistance of the hypothalamus in humans with prediabetes. Treatment with empagliflozin for 8 weeks was able to restore hypothalamic insulin sensitivity, a favorable response that could contribute to the beneficial effects of SGLT2 inhibitors”.
- ***Pituitary insulin resistance***
  5. Pascual-Corrales E, et al. (2024)^13^: Objective: “To investigate the impact of pituitary surgery on glucose metabolism and to identify predictors of remission of diabetes after pituitary surgery in patients with acromegaly”; Study type: National multicenter retrospective study of patients with acromegaly undergoing transsphenoidal surgery; Conclusion: “Glucose metabolism improved in patients with acromegaly after surgery and 21% of the diabetic patients experienced diabetes remission”.
  6. Biagetti B, et al. (2021)^14^; Objective: “To exam whether the Homeostatic Model Assessment of Insulin Resistance is higher in Caucasian, adult, treatment-naïve patients with acromegaly than in the reference population independently of diabetes presence and to evaluate the impact of treatment on its assessment”; Study type: Systematic review and meta-analysis; Conclusion: “The study confirms that insulin resistance is an early event in acromegaly”.
  7. Kinoshita Y, et al. (2011)^15^; Objective: “To identify factors involved in the impairment of glucose metabolism in acromegaly, we evaluated clinical parameters before and immediately after surgical cure of the disease”; Study type: Retrospective study; Conclusion: “Insulin resistance impairs glucose metabolism in acromegaly”.
- ***Thyroid insulin resistance***
  8. Ferrannini E, et al. (2017)^16^; Objective: “To evaluate the relationship between thyroid hormone levels within the normal range and insulin resistance”; Study type: Prospective study and metabolomic analysis; Conclusion: “We demonstrate that serum fT3 concentrations within the euthyroid range are independently associated with insulin resistance both cross-sectionally and longitudinally. This association is supported by a metabolite pattern that points at increased oxidative stress as part of the insulin resistance syndrome”.
  9. Chuang TJ, et al. (2021)^17^; Objective: “To evaluate the relationships between thyroid-stimulating hormone (TSH) and Increased insulin resistance (IR); decreased glucose effectiveness (GE); and both first-and second phase of insulin secretion (FPIS, SPIS) in adult Chinese”; Study type: Cross-sectional study; Conclusion: “The data showed that IR, FPIS, and SPIS were positively related to the TSH level in middle-aged Chinese, whereas GE was negatively related. In both genders, IR had the tightest association followed by GE, FPIS, and SPIS”.
  10. Javed A, et al. (2015)^18^; Objective: “To determine the relationship between TSH concentrations and insulin sensitivity, lipids, and adipokines in euthyroid, non-diabetic, obese *adolescents*”; Study type: Clinical Trial; Conclusion: “Study suggests a sex-specific association between TSH and insulin sensitivity in euthyroid, non-diabetic, obese adolescent males”.
- ***Pulmonary Insulin Resistance***
  11. Sagun G, et al. (2015)^19^; Objective: “To determine if insulin resistance plays a detrimental role in lung function in outpatients admitted to internal medicine clinics in adults from Turkey”; Study type: Cross sectional study; Conclusion: “Insulin resistance should also be considered amongst the contributing factors for decline in lung function”.
  12. Bulcun E, et al. (2012)^20^; Objective: “To investigate the frequency of disorders of glucose metabolism (DGM) and Insulin resistance in patients with obstructive sleep apnoea syndrome and determining factors for these disorders”; Study type: Cross sectional study; Conclusion: “Obstructive sleep apnoea syndrome is associated with high frequency of DGM”.
  13. Huang T, et al. (2022)^21^; Objective: “To examine the risk of developing obstructive sleep apnea (OSA) according to baseline concentrations of fasting insulin and hemoglobin A1c”; Study type: Prospective study; Conclusion: “Independent of obesity, insulin resistance may play a more important role than hyperglycemia in the pathogenesis of OSA”.
  14. Michalek-Zrabkowska M, et al. (2021)^22^; Objective: “The aim of this research was to assess the relationship between prevalence and severity of obstructive sleep apnea (OSA) and insulin resistance among patients with increased risk of OSA without diabetes mellitus”; Study type: Cross sectional study; Conclusion: “Individuals with moderate to severe OSA without diabetes mellitus had a higher prevalence of insulin resistance”.
- ***Myocardial insulin resistance***
  15. Cook SA, et al. (2010)^23^; Objective: “Whole body and myocardial insulin resistance are features of non-insulin-dependent diabetes mellitus (NIDDM) and left-ventricular dysfunction (LVD). We determined whether abnormalities of insulin receptor substrate-1 (IRS1), IRS1-associated PI3K (IRS1-PI3K), and glucose transporter 4 contribute to tissue-specific insulin resistance”; Study type: Analytical study; Conclusion: “The mechanisms of myocardial insulin resistance are different between NIDDM and LVD”.
  16. Iozzo P, et al. (2002)^24^; Objective: “To investigate whether type 2 diabetes is associated with myocardial IR independent of coronary artery disease (CAD)”; Study type: Case-control study; Conclusion: “Type 2 diabetes is specifically associated with myocardial insulin resistance (IR) that is independent of and nonadditive with angiographic CAD and proportional to skeletal muscle and whole-body IR”.
  17. Swan JW, et al. (1997)^25^; Objective: “To assess insulin sensitivity in patients with chronic heart failure (CHF) and its relation to disease severity”; Study type: Cross sectional study; Conclusion: “CHF is associated with marked insulin resistance, characterized by both fasting and stimulated hyperinsulinemia. Advanced heart failure is related to increased insulin resistance, but this is not directly mediated through ventricular dysfunction or increased catecholamine levels”.
  18. Lautamäki R, et al. (2006)^26^; Objective: “To determine the manifestations of metabolic syndrome in different organs in patients with liver steatosis”; Study type: Analytical study; Conclusion: “In patients with type 2 diabetes and coronary artery disease, liver fat content is a novel independent indicator of myocardial insulin resistance and reduced coronary functional capacity”.
- ***Liver insulin resistance***
  19. Lecoultre V, et al. (2014)^27^; Objective: “To assess whether the consumption of chlorogenic acid-rich coffee attenuates the effects of short-term fructose overfeeding, dietary conditions known to increase intrahepatocellular lipids (IHCLs), and blood triglyceride concentrations and to decrease hepatic insulin sensitivity in healthy humans”; Study type: Randomized, controlled, crossover trial; Conclusion: “Coffee consumption attenuates hepatic insulin resistance but not the increase of IHCLs induced by fructose overfeeding. This effect does not appear to be mediated by differences in the caffeine or chlorogenic acid content”.
  20. Fraenkel E, et al. (2023)^28^; Objective: “To assess insulin-like growth factor 1 (IGF-1) and IGF-binding protein 3 (IGFBP3) as markers of insulin resistance in patients with prediabetes and type 2 diabetes mellitus”; Study type: Observational clinical study; Conclusion: “The results demonstrate a fundamental role of IGF-1 and IGFBP3 in the patho-physiology of hepatic insulin resistance and suggest them as indirect indicators of the hepatic insulin resistance”.
  21. Haus JM, et al. (2010)^29^; Objective: “To examine the effects of an exercise/diet lifestyle intervention on free fatty acid (FFA)-induced hepatic insulin resistance in obese humans”; Study type: Clinical Trial; Conclusion: “Both lifestyle interventions are effective in reducing hepatic insulin resistance under basal and hyperinsulinemic conditions. However, the reversal of FFA-induced hepatic insulin resistance is best achieved with a combined exercise/caloric-restriction intervention”.
  22. Miyazaki Y, et al. (2002)^30^; Objective: “To examine the relationship between peripheral/hepatic insulin sensitivity and abdominal superficial/deep subcutaneous fat and intra-abdominal visceral fat in patients with type 2 diabetes mellitus (T2DM)”; Study type: Clinical Trial; Conclusion: “The visceral adiposity is associated with both peripheral and hepatic insulin resistance, independent of gender, in T2DM. In male but not female T2DM, deep subcutaneous adipose tissue also is associated with peripheral and hepatic insulin resistance”.
  23. Kotronen A, et al. (2007)^31^; Objective: “To determine the effect of liver fat on insulin clearance and hepatic insulin sensitivity”; Study type: Clinical Trial; Conclusion: “The increase liver fat is associated with both impaired insulin clearance and hepatic insulin resistance. Hepatic insulin sensitivity associates with liver fat content, independent of insulin clearance”.
- ***Pancreatic insulin resistance***
  24. Wagner R, et al. (2020)^32^; Objective: “To investigate genotype × pancreatic fat interactions on insulin secretion” Study type: Observational study; Conclusion: “The associations suggest that pancreatic steatosis only impairs beta-cell function in subjects at high genetic risk for diabetes. Genetically determined insulin resistance specifically renders pancreatic fat deleterious for insulin secretion”.
  25. Ladwa M, et al. (2021)^33^; Objective: “To compare postprandial insulin secretion and the relationships between insulin secretion, insulin sensitivity and pancreatic fat in men of black West African (BA) and white European (WE) ancestry”; Study type: Cross-sectional study; Conclusion: “Ethnicity is an independent determinant of beta cell function in black and white men. In response to a meal, healthy BA men exhibit lower insulin secretion compared with their WE counterparts for their given insulin sensitivity”.
  26. Weng S, et al. (2018)^34^; Objective: “To explore the prevalence of nonalcoholic fatty pancreas disease (NAFPD) in a Chinese adult population, and investigate factors associated with NAFPD aggravation”; Study type: Cross-sectional study; Conclusion: “The lipid metabolism disorder was the basis for the pathogenesis of NAFPD, and the resulting abnormal secretion of adipokines and ectopic fat deposition in other areas could interact to cause IR and glucose metabolism disorder, which resulted in T2DM”.
  27. Liang B, et al. (2020)^35^; Objective: “To explore the effects of high T3 levels on β-cell line insulin resistance, as well as the roles of endoplasmic reticulum stress (ERS)”; Study type: Analytical study; Conclusion: “High T3 levels can induce insulin resistance in β-cell line by activating ERS and the apoptotic pathway”.
- ***Renal insulin resistance***
  28. Becker B, et al. (2005)^36^; Objective: “The relationship among insulin resistance, adiponectin, and cardiovascular (CV) morbidity in patients with mild and moderate kidney disease was investigated”; Study type: Prospective study; Conclusion: “In patients with chronic kidney diseases, a syndrome of insulin resistance is present even in the earliest stage of renal dysfunction, and several components of this syndrome are associated with CV events”.
  29. Landau M, et al. (2011)^37^; Objective: “ To assess whether the factors associated with insulin resistance (IR) were different in those with and without chronic kidney disease (CKD)”; Study type: Analytical study; Conclusion: “In Stage 3 CKD, kidney function is associated with IR; except for adiponectin, the correlates of IR are similar in those with and without CKD”.
- ***Spleen insulin resistance***
  30. El-Aziza RA, et al. (2018)^38^; Objective: “To assess spleen longitudinal diameter (SLD) in patients with metabolic syndrome (MS) and to investigate the possible factors affecting spleen size”; Study type: Case-control study; Conclusion: “Patients with MS had larger spleen size than healthy controls. SLD significantly correlated with waist circumference but not IL-10 in patients with MS”.
- ***Small intestine Insulin Resistance***
  31. Urita Y, et al. (2006)^39^; Objective: “To investigate non-invasively the incidence of absorption of carbohydrates in diabetic patients during an oral glucose tolerance test and to determine whether malabsorption may be associated with insulin secretion and insulin resistance”; Study type: Cross-sectional study; Conclusion: “Insulin resistance may be overestimated by using these markers if the patient has carbohydrate malabsorption, or that carbohydrate malabsorption may be present prior to the development of insulin resistance. Hence carbohydrate malabsorption should be taken into account for estimating insulin resistance and beta-cell function”.
  32. Angelini G, et al. (2021)^40^; Objective: “To assess the role of jejunum in insulin resistance in humans and in experimental animals”; Study type: Observational study; Conclusion: “Proximal gut plays a crucial role in controlling insulin sensitivity through a distinctive metabolic signature involving hepatic gluconeogenesis and muscle insulin resistance. Bypassing the jejunum is beneficial in terms of insulin-mediated glucose disposal in obesity”.
  33. Lalande C, et al. (2020)^41^; Objective: “To evaluate small intestine epithelial cell homeostasis in a cohort of men covering a wide range of adiposity and glucose homoeostasis statuses”; Study type: Cross-sectional study; Conclusion: “A decreased functional enterocyte mass and an increased enterocyte death rate in presence of metabolic alterations but emphasizes that epithelial cell homeostasis is especially altered in presence of severe insulin resistance and T2D. The marked changes in small intestine cellularity observed in obesity and diabetes are thus suggested to be part of gut dysfunctions, mainly at an advanced stage of the disease”.
- ***Large intestine insulin resistance***
  34. Honka H, et al. (2013)^42^; Objective: “to validate, using an animal model, the use of positron emission tomography (PET) in the estimation of intestinal glucose uptake (GU), and thereafter to test whether intestinal insulin-stimulated GU is altered in morbidly obese compared with healthy human participants”; Study type: Clinical study; Conclusion: “Intestinal GU can be quantified in vivo by [(18)F]FDG PET. Intestinal insulin resistance occurs in obesity before the deterioration of systemic glucose tolerance”.
  35. Gao C, et al. (2022)^43^; Objective: “To investigate the effects of intestinal alkaline phosphatase (IAP) in controlled intestinal inflammation and alleviated associated insulin resistance (IR)”; Study type: In vitro study; Conclusion: “The IAP can be used as a natural anti-inflammatory agent to reduce intestinal inflammation-induced IR”.
- ***Muscle insulin resistance***
  36. Magkos F, et al. (2012)^44^; Objective: “To evaluate the relationship between the rate of release of free fatty acids (FFA) into plasma and skeletal muscle insulin sensitivity in human subjects”; Study type: Clinical Trial; Conclusion: “The data suggest that the correlation between FFA kinetics and muscle glucose metabolism is due to multiorgan insulin resistance rather than a direct effect of FFA itself on skeletal muscle insulin action and challenge the view that increased adipose tissue lipolytic rate is an important cause of insulin resistance”.
  37. Nowotny B, et al. (2013)^45^; Objective: “To examine initial events occurring during the onset of insulin resistance upon oral high-fat loading compared with lipid and low-dose endotoxin infusion”; Study type: Clinical Trial; Conclusion: “The oral fat ingestion rapidly induces insulin resistance by reducing nonoxidative glucose disposal, which associates with muscle PKCθ activation and a rise in distinct myocellular membrane diacylglycerols, while endotoxin-induced insulin resistance is exclusively associated with stimulation of inflammatory pathways”.
  38. Abbasi F, et al. (2000)^46^; Objective: “To evaluate the ability of insulin to regulate free fatty acid (FFA) concentrations in healthy nondiabetic subjects selected to be either insulin-resistant or -sensitive on the basis of insulin-mediated glucose disposal by muscle”; Study type: Clinical Trial; Conclusion: “The results *demonstrate* that the ability of insulin to regulate plasma FFA concentrations is impaired in healthy subjects with muscle insulin resistance, indicating that insulin-resistant individuals share defects in the ability of insulin to stimulate muscle glucose disposal and to inhibit adipose tissue lipolysis”.
  39. Krebs M, et al. (2002)^47^; Objective: “To examine effects of short-term plasma amino acid (AA) elevation on whole-body glucose disposal and cellular insulin action in skeletal muscle”; Study type: Clinical Trial; Conclusion: “The plasma amino acid elevation induces skeletal muscle insulin resistance in humans by inhibition of glucose transport/phosphorylation, resulting in marked reduction of glycogen synthesis”.
  40. Kelley DE, et al. (2001)^48^; Objective: “To examine the respective roles of plasma free fatty acids, regional adiposity, and other metabolic factors as determinants of the severity of skeletal muscle insulin resistance (IR) in type 2 diabetes mellitus (DM)”; Study type: Clinical Trial; Conclusion: “The severity of skeletal muscle IR in type 2 DM is closely related to the IR of suppressing lipolysis and that plasma fatty acids and visceral adipose tissue are key elements mediating the link between obesity and skeletal muscle IR in type 2 DM”.
- ***Adipose Tissue insulin resistance***
  41. Halloun R, et al. (2023)^49^; Objective: “To teste whether adipose tissue insulin sensitivity predicts changes in the degree of obesity over time” Study type: Secondary analysis of an observational study; Conclusion: “The adipose tissue insulin resistance is not protective from increases of the degree of obesity and skeletal muscle insulin resistance is not associated with increases of the degree of obesity”.
  42. Hazlehurst JM, et al. (2013)^50^; Objective: “To determine whether glucocorticoids have tissue-specific effects on insulin sensitivity in vivo”; Study type: Double-blind, randomized, placebo-controlled, crossover study; Conclusion: “The human subcutaneous adipose insulin sensitization by glucocorticoids in vivo demonstrates tissue-specific actions of glucocorticoids to modify insulin action”.
  43. Cifarelli V, et al. (2020)^51^; Objective: “To evaluate the potential influence of adipose tissue (AT) oxygenation on AT biology and insulin sensitivity in people”; Study type: Clinical Trial; Conclusion: “To reduce AT oxygenation in individuals with obesity contributes to insulin resistance by increasing plasma PAI-1 concentrations and decreasing AT branched-chain amino acid (BCAA) catabolism and thereby increasing plasma BCAA concentrations”.
  44. Jiang J, et al. (2020)^52^; Objective: “To examine the association of different anatomical forms of obesity with adipose tissue insulin resistance and to assess the diagnostic value and contribution of obesity to adipose tissue insulin resistance”; Study type: Cross-sectional study; Conclusion: “Maintaining waist circumference in males and body mass index in females to a normal range could be an important strategy to significantly reduce the occurrence of adipose tissue insulin resistance and the subsequent metabolic diseases”.
  45. Ter Horst KW, et al. (2017)^53^; Objective: “To validate simplified methods for the quantification of adipose tissue insulin resistance against the assessment of insulin sensitivity of lipolysis suppression during hyperinsulinemic-euglycemic clamp studies”; Study type: Analytical study; Conclusion: “Adipose tissue insulin sensitivity can be reliably quantified in overweight and obese humans by simplified index methods. The sensitivity and specificity of the Adipo-IR index and the fasting plasma insulin-glycerol product, combined with their simplicity and acceptable agreement, suggest that these may be most useful in clinical practice”.
  46. Wen J, et al. (2020)^54^; Objective: “The degree of adipose tissue insulin resistance increases in obesity, prediabetes and type 2 diabetes, but whether it associates with prediabetes is unclear”; Study type: Cross-sectional study; Conclusion: “Adipose tissue insulin resistance is associated with prediabetes and should be considered for use in population studies”.
  47. Zhou Q, et al. (2024)^55^; Objective: “To examine the association between adipose tissue-specific insulin resistance and atherosclerotic burden and plaques in intracranial, extracranial, and coronary arteries in community residents without diabetes”; Study type: Clinical Trial; Conclusion: “Adipose tissue-specific insulin resistance is associated with atherosclerotic burden and plaques in intracranial and coronary arteries in Chinese community nondiabetic residents”.
- ***Vascular insulin resistance***
  48. Feldman RD, et al. (1996)^56^; Objective: “To determine whether dietary salt restriction might affect vascular sensitivity to insulin”; Study type: Prospective study; Conclusion: “In these younger normotensive and hypertensive subjects, dietary salt restriction increases resistance to the vasodilating effects of insulin”.
  49. Wang N, et al. (2020)^57^; Objective: “To examine whether GLP-1 recruits microvasculature and improves the action of insulin in obese humans”; Study type: Clinical Trial; Conclusion: “In obese humans with microvascular insulin resistance, GLP-1’s vasodilatory actions are preserved in both skeletal and cardiac muscle microvasculature, which may contribute to improving metabolic insulin responses and cardiovascular outcomes”.
  50. Vinet A, et al. (2015)^58^; Objective: “To assess the insulin vasoreactivity in metabolic syndrome (MetS), and to evaluate the effects of a lifestyle program”; Study type: Case-control study; Conclusion: “The local vasodilatory effects to insulin and its overall flow motion are impaired in MetS subjects in relation to inflammation. The lifestyle intervention reversed this insulin-induced vascular dysfunction in parallel to decreased inflammation level”.
  51. Love KM, et al. (2024)^59^; Objective: “To elucidate the impact of elevated free fatty acids (FFAs) on insulin action across the arterial tree and define the relationship among insulin actions in the different arterial segments”; Study type: Randomized crossover study; Conclusion: “Clinically relevant elevation of plasma FFA concentrations induces pan-arterial insulin resistance, the vascular insulin resistance outcomes are interconnected, and insulin-mediated muscle microvascular perfusion associates with cardiovascular disease predictors. Our data provide biologic plausibility whereby a causative relationship between FFAs and cardiovascular disease could exist, and suggest that further attention to interventions that block FFA-mediated vascular insulin resistance may be warranted”.
- ***Ovarian insulin resistance***
  52. Wu XK, et al. (2003)^60^; Objective: “Insulin resistance is a common feature of both polycystic ovary syndrome (PCOS) and non-insulin-dependent diabetes mellitus (NIDDM); however, the persistent reproductive disturbances appear to be limited to the former, suggesting that insulin resistance in the ovary itself may confer this susceptibility” Study type: Prospective study; Conclusion: “There is a selective defect in insulin actions in PCOS granulosa cells, which suggests ovarian insulin resistance, and this metabolic phenotype is associated with an enhanced IGF-1 mitogenic potential. Troglitazone could divergently alter expression of various IRS molecules and insulin actions and could be used as an ovarian insulin sensitizer and mitogen/steroidogenic inhibitor in PCOS”.
- Testicular insulin resistance
  53. Contreras PH, et al. (2018)^61^; Objective: “To evaluate insulin sensitivity and testicular function in a cohort of adult males suspected of being insulin-resistant”; Study type: Prospective study; Conclusion: “Waist Circumference predicted both insulin resistance (>99 cm) and hypogonadism (>110 cm), suggesting that the first hit of abdominal obesity is insulin resistance and the second hit is male hypogonadism. Normal weight did not protect from insulin resistance, while a relevant proportion of obese subjects were non-insulin resistant”.
  54. Verit A, et al. (2014)^62^; Objective: “To investigate the possible effect of insulin resistance (IR) on male reproductive system via evaluation of semen analysis, male sex hormones and serum lipid profiles, and testicular volumes”; Study type: Prospective study; Conclusion: “IR may be accused of causing detrimental effect on male infertility due to hyperinsulinemic state and being one of the components for MetS. Interestingly, due to our preliminary results, we do not found any inverse correlation between IR and male reproductive functions”.

## INTERPRETATION OF FINDINGS

Given the novelty of the term “In Situ Diabetes Mellitus” and the proposed “Type 6 Diabetes Mellitus,” it was imperative to ensure that the objectives of our study aligned with the existing body of diabetes research, clinical practices, and potential implications. Our concept of T6DM refers to the localized development of IR in specific organs, such as the brain, hypothalamus, pituitary gland, thyroid, lung, heart, liver, pancreas, kidneys, spleen, small intestine, large intestine, muscle, adipose tissue, vessels, ovaries, and testicles, independent of the systemic insulin sensitivity. We evaluated organ- and system-specific IR, which was characterized as T6DM. The scoping review presented in this study provides an overview of the existing literature on organ-specific IR, a condition characterized by the development of IR in individual organs independent of systemic metabolic disturbances. Our results highlight the complex nature of this condition, involving a wide range of tissues and organs.

IR manifests through various mechanisms that differ across tissues, yet several critical signaling pathways are frequently impaired. These impairments often involve a reduction in the activity of insulin receptor tyrosine kinases, changes in the phosphorylation of IRS, and the dysregulation of downstream signaling components, including phosphatidylinositol 3-kinase (PI3K), and protein kinase B (Akt). Additionally, chronic inflammation may worsen IR by activating inflammatory signaling pathways, particularly those associated with tumor necrosis factor alpha (TNF-α) and nuclear factor-κB. In this regard, each organ or tissue can be considered a potential locus of IR, with the degree of severity and the specific molecular mechanisms influenced by factors such as genetic predisposition, environmental conditions, and the presence of comorbidities.

### Brain in situ diabetes mellitus

The traditional view of the brain as insulin-impervious has been challenged by the discovery of insulin receptors in various brain regions. These findings highlight the role of insulin in modulating neuronal function. Disruptions in insulin signaling have been implicated in Alzheimer’s disease (AD).^63^ Brain-IR is characterized by decreased neuronal responsiveness to insulin, leading to impaired metabolic function and cognitive processes. This attenuated response can result from reduced insulin receptor expression, impaired insulin binding, or defects in insulin signaling. These alterations disrupt insulin-mediated cerebral physiology, leading to neurotransmitter dysfunction and impaired neuroplasticity.^64^ The transport of insulin across the blood-brain barrier is influenced by several factors, including blood-brain barrier permeability, genetics, age, body mass index, and blood glucose and triglyceride levels.^65^ While the brain was previously thought to rely solely on insulin-independent glucose transporters, recent research has suggested that it may synthesize its own insulin.^66^ However, the pancreas is the primary source of circulating insulin.^67^

Chronic peripheral hyperinsulinemia can lead to decreased insulin receptor density at the blood-brain barrier, resulting in diminished insulin signaling in the brain.^68^ Additionally, age-related reductions in brain insulin receptors have been implicated in the development of brain-IR. Although brain-IR is thought to be independent of T2DM, it can contribute to alterations in synaptic plasticity, beta-amyloid accumulation, and cognitive impairment.^69^

Insulin exerts a neuroprotective and trophic effect on cerebral cells, attenuating apoptosis, oxidative stress, and beta-amyloid toxicity. Brain-IR and impaired cerebral glucose metabolism are consistently observed in AD and other neurodegenerative disorders.^70^ Systemic inflammation associated with obesity can induce brain-IR, manifesting in various ways depending on the affected brain region. Individuals with age-related cognitive decline or AD often exhibit signs of brain-IR, including elevated insulin receptor serine-1 substrate phosphorylation and decreased cerebrospinal fluid insulin concentrations.^71^

Based on the evidence presented, it is evident that brain-IR closely aligns with the pathophysiological characteristics of T2DM, suggesting a potential analogy to “brain in situ diabetes mellitus.” The complex interplay between insulin signaling, mitochondrial function, and inflammation within the brain underscores the importance of targeting these pathways for therapeutic interventions aimed at attenuate the cognitive decline associated with neurodegenerative diseases.

### Hypothalamus in situ diabetes mellitus

Hypothalamic-IR represents a unique metabolic alteration characterized by a selective impairment of insulin signaling within the hypothalamus. Unlike peripheral IR, hypothalamic-IR disrupts the intricate regulatory mechanisms controlling energy homeostasis, leading to a cascade of metabolic dysfunctions. This condition is implicated in the development of obesity and T2DM, highlighting the critical role of hypothalamic insulin sensitivity in maintaining metabolic equilibrium.^72^

Within the hypothalamus, insulin plays a fundamental role in regulating both food intake and glucose metabolism, and dysregulation of these processes is intimately linked to the development of obesity and T2DM.^73^ The pathogenesis of hypothalamic-IR is a multifaceted process involving several interconnected mechanisms. Chronic inflammatory responses in the hypothalamus, frequently triggered by high-fat diets or genetic predispositions, can change insulin signaling pathways. Excessive nutrient intake and lipid accumulation within hypothalamic cells can disrupt insulin receptor function and downstream signaling cascades.^74^ The pathophysiology of hypothalamic-IR is characterized by complex interactions between various pathways and factors. Endoplasmic reticulum stress-induced activation of the Activating Transcription Factor 4-Serine 6 kinase pathway and decreased serine phosphorylation of insulin receptor substrate-1 (IRS-1) have been implicated in the development of this metabolic condition.^75^

The therapeutic setting for hypothalamic-IR remains a dynamic area of investigation, with ongoing efforts to identify novel targets and therapeutic strategies. Several promising avenues are currently under exploration. One area of particular interest is targeting inflammatory pathways. Chronic low-grade inflammation, a hallmark of HIR, can be modulated through interventions aimed at pro-inflammatory cytokines and immune mediators. This approach offers potential for novel pharmacological interventions. Additionally, the gut microbiome has emerged as a key player in metabolic regulation. Alterations in gut microbiota composition have been associated with hypothalamic-IR, suggesting that therapies targeting the microbiome could potentially modulate hypothalamic insulin sensitivity. Finally, combination therapies, utilizing multiple pharmacological agents with complementary mechanisms of action, may offer synergistic benefits in improving insulin sensitivity and metabolic effects.^76^

While the term “hypothalamus in situ diabetes mellitus” is not a universally recognized medical term, it could be interpreted as describing a state where IR specifically within the hypothalamus leads to metabolic dysregulation similar to that observed in systemic DM. This condition would imply that the hypothalamus itself would be experiencing a form similar to T2DM, characterized by impaired glucose uptake and utilization.

### Pituitary in situ diabetes mellitus

Evidence supports the hypothesis that IR can manifest in the pituitary gland, leading to impaired insulin signaling and subsequent hormonal dysregulation. This understudied phenomenon, termed pituitary-IR, has been the subject of increasing research interest within the field of endocrinology. Several mechanisms have been proposed to explain the development of pituitary-IR. One potential pathway involves the activation of inflammatory processes within the pituitary gland, leading to the overproduction of reactive oxygen species, exacerbating oxidative stress. Consequently, IR develops, and the pituitary gland’s ability to synthesize and release hormones is compromised.^77^

Pituitary-IR is a complex disorder characterized by impaired insulin signaling within the pituitary gland, leading to disrupted glucose homeostasis in pituitary gland. The precise mechanisms underlying pituitary-IR remain an active area of investigation, but accumulating evidence suggests that this condition involves a multifaceted interplay of factors.^78^ Downregulation of insulin receptors and dysregulation of post-receptor signaling pathways, such as the PI3K/Akt pathway, have been implicated in the pathogenesis of pituitary-IR. Furthermore, IRS proteins, critical for glucose uptake and metabolism, may be impaired in this condition.^79^ Chronic hyperinsulinemia, often associated with obesity, can exacerbate pituitary-IR by inducing negative feedback mechanisms that further diminish insulin sensitivity. Additionally, inflammatory cytokines and increased free fatty acids contribute to the dysfunction of insulin signaling within the pituitary gland.^80^

Diagnosing pituitary-IR remains a clinical challenge due to the complex interplay of endocrine and metabolic factors. While traditional methods for assessing insulin sensitivity, such as the homeostatic model assessment of IR (HOMA-IR), can provide preliminary insights, they may not be entirely specific for pituitary-IR. Direct assessment of insulin signaling within the pituitary gland is often impractical in clinical settings. Consequently, a comprehensive diagnostic approach typically involves a combination of clinical evaluation, biochemical tests, and imaging studies. Measurement of pituitary hormones, including growth hormone and cortisol, can provide valuable clues about the underlying pathophysiology. Dynamic tests such as the oral glucose tolerance test and the insulin suppression test may be helpful in characterizing insulin sensitivity and secretion. Furthermore, molecular studies focusing on the genetic factors influencing insulin signaling pathways are essential for a comprehensive understanding of this condition. However, the gold standard for diagnosing pituitary-IR remains elusive and further research is needed to develop more specific and sensitive biomarkers.^81,82^

Despite the established efficacy of pharmacological interventions in managing peripheral IR, their application to pituitary-IR is less clear. Although these medications may offer potential therapeutic avenues, there is a paucity of data regarding their efficacy and safety in targeting IR specifically within the pituitary gland. Therapeutic interventions for pituitary-IR are currently limited and often tailored to the underlying etiology. Lifestyle modifications, including weight loss and regular physical activity, are foundational to improving insulin sensitivity. Pharmacologic approaches, such as metformin and thiazolidinedione, have shown promise in ameliorating IR in peripheral tissues but their efficacy in pituitary-IR is less established.^83^ Given the complex interplay of endocrine factors contributing to pituitary-IR, targeted therapies aimed at specific molecular pathways are an area of research. Thus, agents that modulate the hypothalamic-pituitary axis or directly target insulin signaling within the pituitary may offer novel therapeutic avenues.

Thus, the phenomenon of pituitary-IR, which could be conceptualized as “pituitary in situ diabetes mellitus”, represents a substantial research frontier within the field of endocrinology. Despite the inherent challenges associated with diagnosis and treatment, a growing body of evidence underscores the intricate interplay of factors contributing to this condition. Future research endeavors should focus on elucidating the precise molecular mechanisms underlying pituitary-IR, developing more sensitive and specific biomarkers for its diagnosis, and exploring novel therapeutic strategies that target the specific molecular pathways involved in the pathogenesis of this disorder. Such advancements are essential for improving the management of pituitary-IR and fostering a deeper understanding of the complex interplay between insulin signaling and endocrine function.

### Thyroid in situ diabetes mellitus

Studies have highlighted the complex interplay between thyroid hormones and insulin signaling pathways, suggesting that thyroid dysfunction can exacerbate insulin resistance.^84^ This relationship have demonstrated that thyroid dysfunction can contribute to IR, highlighting the complex interplay between these endocrine systems.^85^

The molecular mechanisms underlying thyroid-IR are multifaceted. One of the key mechanisms linking thyroid dysfunction to IR involves the modulation of glucose transporter expression. Thyroid hormones are known to regulate the expression of glucose transporter type 4 (GLUT4), a critical glucose transporter in adipose tissue and muscle.^86^ In hypothyroid states, the downregulation of GLUT4 leads to decreased glucose uptake, contributing to hyperglycemia and IR. Additionally, thyroid hormones influence the activity of key enzymes involved in glucose metabolism, further complicating the metabolic landscape in thyroid-IR.^87^

Fluctuations in thyroid hormones concentrations can potentially modulate insulin signaling cascades, culminating in IR.^88^ Moreover, IR can also exert a reciprocal influence on thyroid function. Insulin plays an important role in the transformation of thyroid hormones from their inactive precursor, thyroxine (T4), into their biologically active form, triiodothyronine (T3), within peripheral tissues. IR interferes with this metabolic process, leading to a reduction in T3 production and an elevation in reverse T3, an inactive thyroid hormone derivative.^84^ A state of thyroid hormone imbalance can contribute to the development of hypothyroidism.

Hypothyroidism is a significant contributing factor to IR. Reduced thyroid hormone concentrations are associated with impaired glucose disposal in skeletal muscle, a consequence of diminished insulin sensitivity. Furthermore, hepatic gluconeogenesis is upregulated in hypothyroidism, further exacerbating IR. These physiological alterations underscore the intricate relationship between thyroid dysfunction and metabolic disorders.^89^

Thyroid hormones actively participate in glucose homeostasis, promoting hepatic glycogen synthesis and glucose production. Furthermore, these hormones regulate the expression of key enzymes involved in peripheral glucose uptake and metabolism. Clinical investigations have consistently revealed a greater prevalence of thyroid dysfunction in diabetic patients relative to the general population.^90^

The pathophysiological link between IR and thyroid dysfunction is increasingly evident, especially in obese individuals. Research has consistently demonstrated a strong correlation between IR and the prevalence of thyroid nodules.^91^

Thus, the intricate interplay between thyroid hormones and insulin signaling pathways underscores the critical role of thyroid function in glucose metabolism. Thyroid dysfunction can exacerbate IR through mechanisms involving glucose transporter regulation, enzyme activity modulation, and insulin signaling interference. Conversely, IR can also impair thyroid hormone production, creating a vicious cycle that contributes to metabolic disorders. These findings highlight the importance of comprehensive assessment and management of thyroid function in individuals with IR, particularly those with obesity and thyroid nodules.

### Pulmonary in situ diabetes mellitus

Pulmonary-IR, a condition characterized by the diminished sensitivity of lung tissues to insulin, has emerged as a focal point of scientific inquiry. While the presence of insulin receptors in the lungs underscores the direct influence of insulin on pulmonary function, the mechanisms underlying pulmonary-IR remain elusive.^9392^ Chronic inflammation, a well-established contributor to IR in other organs, is believed to play an important role in the development of pulmonary-IR. The release of pro-inflammatory cytokines and other inflammatory mediators can disrupt insulin signaling pathways, leading to resistance. Additionally, oxidative stress, characterized by an increased production of reactive oxygen species, has been implicated in the pathogenesis of pulmonary-IR. Oxidative stress can impair insulin signaling and lung function, and it may be exacerbated by exposure to pollutants and smoking, both of which are known risk factors for respiratory diseases.^93^

The identification of insulin receptors in pulmonary tissue has showed a previously underappreciated link between metabolic health and respiratory function. Impaired insulin signaling, as observed in IR, can adversely affect lung structure and function. Consequently, IR emerges as a standalone risk factor for the development of respiratory diseases.^94^

IR has been implicated in the development of pulmonary dysfunction through a complex interplay of mechanisms. IGF-1, a key mediator of insulin signaling, can influence the contractility of airway smooth muscle by stimulating its proliferation. Additionally, studies have revealed a correlation between severe asthma and leptin resistance, suggesting a potential link between metabolic disorders and respiratory conditions.^95^ Moreover, adiponectin and cytokines, which are closely associated with IR, exhibit pro-inflammatory effects in the lungs by promoting fibroblast production. Furthermore, a strong correlation exists between IR and bronchial hyperresponsiveness, a hallmark of asthma and other respiratory diseases. Animal models have shed light on the underlying mechanisms, demonstrating that airway inflammation can impair insulin receptor signaling, leading to a cascade of events that promote a pro-inflammatory response and exacerbate IR.^96^

Thus, the evidence presented underscores the relationship between IR and pulmonary function. The identification of insulin receptors in the lungs, coupled with the elucidation of mechanisms such as inflammation and oxidative stress, establishes IR as a significant contributor to pulmonary dysfunction. These findings highlight the need for a more integrated approach to the management of respiratory diseases, considering both metabolic and pulmonary factors. The concept of “pulmonary in situ diabetes mellitus” offers a framework for understanding the complex interplay between IR and pulmonary function.

### Heart in situ diabetes mellitus

Evidence suggests that the heart may serve as both a marker and a target of systemic IR. The concept of myocardial-IR posits that cardiac tissue can develop IR independently, contributing directly to cardiac disease.^97^ Myocardial-IR is functionally defined by impaired glucose uptake and metabolism within the myocardium. While myocardial-IR frequently accompanies systemic IR, the precise mechanisms underlying this association remain an active area of investigation.^98^

The heart is a responsive tissue to insulin action. Studies have consistently demonstrated a high prevalence of myocardial-IR among patients with heart failure. Curiously, this association persists even in diabetic patients with well-controlled blood glucose levels, implying a direct causal relationship between myocardial-IR and the development of heart failure.^99^

The intracellular mechanisms governing myocardial-IR are complex and multifaceted. Specific signaling pathways within the myocardium have been shown to be activated in individuals with systemic IR.^100^ Myocardial-IR is characterized by a disruption in the normal insulin signaling pathways, leading to an elevated risk of cardiac dysfunction. It is essential to differentiate between the effects of hyperactive signaling pathways that remain responsive to insulin and those caused by impaired insulin-mediated glucose regulation. The consequences of altered insulin signaling in cardiac muscle are many-sided and are closely associated with systemic metabolic dysfunction.^101^

### Liver in situ diabetes mellitus

Hepatic-IR stands as an important factor in the pathogenesis of type T2DM and a constellation of metabolic disorders. This complex phenotype arises from a confluence of genetic, environmental, and lifestyle determinants.^102^ Hepatic-IR is characterized by a blunted response to insulin, manifesting as impaired hepatic glucose uptake and increased hepatic glucose output. Concomitantly, alterations in hepatic lipid metabolism contribute to the development of this metabolic derangement. Several molecular mediators intracellular and signaling cascades have been implicated in the pathogenesis of Hepatic-IR. Prominent among these are serine/threonine kinases, including protein kinase C-epsilon and c-Jun N-terminal kinase.^103^ These kinases play a pivotal role in propagating inflammatory signals and inducing cellular stress. Moreover, perturbations in endoplasmic reticulum homeostasis and oxidative stress contribute to the development of Hepatic-IR.^104^ Collectively, these factors converge to impair insulin signaling and promote hepatic glucose production and lipogenesis.

Hepatic-IR profoundly impacts systemic glucose regulation and lipid handling. Under physiological conditions, insulin exerts an inhibitory effect on hepatic glucose output while stimulating glycogen storage. Conversely, in the state of hepatic-IR, the liver exhibits a paradoxical increase in glucose production despite elevated circulating insulin concentrations. This aberrant hepatic glucose flux contributes to hyperglycemia and perpetuates systemic IR.^105^ Moreover, hepatic-IR is characterized by enhanced hepatic lipogenesis and impaired triglyceride clearance, culminating in hepatic steatosis and dyslipidemia. Considering the significant metabolic consequences of hepatic-IR, numerous therapeutic interventions have been investigated with the aim of ameliorating hepatic insulin sensitivity, suppressing hepatic glucose production, and restoring normolipidemia.^106^

Therefore, hepatic-IR emerges as a complex pathophysiological process that is central to the development of T2DM and associated metabolic disturbances. A profound understanding of the underlying mechanisms driving hepatic-IR is imperative for the design of efficacious therapeutic interventions. While lifestyle modifications and conventional pharmacological agents provide promising avenues for managing this condition, ongoing research endeavors hold the potential to unveil novel and more precise therapeutic modalities. By comprehensively addressing hepatic-IR, we can significantly enhance metabolic health and alleviate the burden imposed by T2DM.

### Pancreas in situ diabetes mellitus

Pancreatic-IR is a many-sided physiological occurrence that holds substantial importance in the progression of T2DM. Pancreatic-IR denotes a condition in which there is a disruption in insulin secretion by the pancreatic β-cells. This disruption in insulin secretion often happens concurrently with peripheral IR, establishing a detrimental cycle that worsens the advancement of the disease.^107^

The mechanisms underlying pancreatic-IR are complicated and heterogeneous, encompassing a complex interplay of genetic, hormonal, and environmental factors. Genetic predisposition plays a substantial role in the onset of pancreatic IR by impacting the structural and functional integrity of pancreatic β-cells. The identification of specific genetic variants has provided insights into the molecular mechanisms underlying the increased susceptibility of certain individuals to both pancreatic-IR and T2DM.^108^

Pancreatic-IR is a critical factor in the pathogenesis of T2DM. Hormonal imbalances, including elevated levels of free fatty acids and adipokines, can impair the function of pancreatic β-cells and promote insulin resistance. The chronic inflammation can further exacerbate pancreatic-IR, and this has significant implications for overall metabolic health. Although peripheral IR plays a role in the development of T2DM, pancreatic-IR worsens the progression of the disease by hindering β-cell function and insulin secretion.^109^

A comprehensive understanding of the underlying mechanisms and clinical consequences of pancreatic-IR is indispensable for the development of efficacious therapeutic interventions. The insufficiency of insulin secretion, often attributed to β-cell dysfunction, represents a critical juncture in the progression of this metabolic disorder.

### Kidney in situ diabetes mellitus

Insulin signaling is a complex process divided into primary pathways, including the PI3K/Akt and MAPK/MEK cascades. In IR, these pathways are not uniformly affected. Recent evidence suggests that selective impairment of insulin signaling pathways in the kidney also contributes to this condition.^101^

The kidney possesses the unique ability to generate glucose through renal neoglycogenesis. While a significant portion of this glucose is utilized by the kidney itself, excessive renal glucose production can contribute to hyperglycemia. In the renal proximal tubule, insulin signaling mediated by IRS1 is suppressed, whereas IRS2-dependent signaling persists. The deficiency of IRS1 signaling in the proximal tubule may compromise IRS1-mediated inhibition of gluconeogenesis, potentially exacerbating hyperglycemia by sustaining glucose production. In the glomerulus, impaired IRS1 signaling can adversely affect the structure and function of podocytes and endothelial cells, potentially contributing to the development of diabetic nephropathy.^111^

The development of renal-IR is influenced by several interconnected elements. Persistent elevated blood glucose levels, a characteristic of DM, are important in initiating renal-IR by inducing oxidative stress and inflammatory responses within kidney tissues. Furthermore, the progression and onset of renal-IR are exacerbated by abnormalities in the renin-angiotensin-aldosterone system, heightened activity of the sympathetic nervous system, and increased formation of advanced glycation end-products.^112^ The clinical significance of renal-IR extends beyond DM management. Research indicates that renal-IR plays an important role in the development of diabetic nephropathy, which is a primary contributor to end-stage renal disease globally. Furthermore, renal-IR is linked to ongoing kidney function decline, elevated blood pressure, and heart-related complications in both diabetic and non-diabetic populations.^113^

Although substantial progress has been achieved in unraveling the complexities of the existence of selective IR in the kidney, numerous questions remain unanswered. By delving into the cellular and molecular alterations associated with this condition, we can gain invaluable insights into its pathogenesis.

### Spleen in situ diabetes mellitus

The spleen plays an important role in the induction or blockade of IR, as monocytes, once secreted by the bone marrow, do not enter circulation immediately but are stored in the spleen.^114^ During inflammatory events, including those occurring in adipose tissue, monocytes migrate to the site of inflammation and differentiate into pro-inflammatory macrophages, thereby contributing to the development of IR.^115^

Studies have shown that splenectomized individuals with obesity are protected from IR.^116^ This is likely due to the reduced availability of monocytes to differentiate into inflammatory macrophages in adipose tissue, leading to preserved insulin signaling pathways and reduced inflammatory responses.

Consuming an excess of calories sets off inflammatory signaling pathways, induces stress in the endoplasmic reticulum, and stimulates Toll-like receptor 4, resulting in IR within the hypothalamus, muscle tissues, and liver. Simultaneously, the volume of adipose tissue expands, drawing in monocytes that penetrate the fat tissue and undergo differentiation. As a result, the onset of IR becomes evident; however, the worsening of IR in obese people is dependent on mechanisms mediated by the spleen, where monocytes transform into macrophages, further contributing to the development of IR.^117^

The complex relationship between physiological and pathological factors in spleen-IR underscores the spleen’s important function in maintaining systemic glucose balance. Emerging research indicates that spleen-IR can result from various conditions, such as obesity, T2DM, and persistent inflammation. The precise mechanisms underlying spleen-IR are still being investigated, researchers hypothesize that disrupted insulin signaling in splenic cells, especially macrophages, plays a role in the onset of systemic IR. Additionally, spleen-IR may intensify inflammatory processes within the organ, creating a self-perpetuating cycle that worsens metabolic dysfunction.

### Small intestine in situ diabetes mellitus

Small intestine-IR is a complex metabolic disorder characterized by compromised insulin signaling and glucose uptake, resulting in disrupted nutrient absorption and energy balance. While research has been conducted on IR in peripheral tissues, the underlying mechanisms of small intestinal-IR remain largely unclear. Findings indicate that changes in gut microbiome composition, inflammatory processes, and genetic components may play a role in the onset of IR within the small intestine. Understanding the molecular mechanisms involved in small intestinal-IR is important for the development of targeted treatments aimed at enhancing metabolic health and preventing the progression to T2DM.

The small intestine plays a fundamental role in glucose homeostasis, particularly in modulating insulin sensitivity. Upon food intake, the small intestine releases incretin hormones, including glucose-dependent insulinotropic polypeptide (GIP) and glucagon-like peptide-1 (GLP-1).^118^ These hormones significantly enhance insulin secretion from pancreatic β-cells, increasing it by more than 50% in response to a meal.^119^ Beyond their pancreatic actions, GIP and GLP-1 exert other effects on various tissues, including the central nervous system. By acting on specific neuronal circuits, these incretins contribute to reduced food intake and increased energy expenditure.^120^ Additionally, they possess insulinomimetic actions, enhancing glucose uptake by peripheral tissues like skeletal muscle and suppressing hepatic glucose production.^121^

The small intestine’s primary role includes the effective uptake and processing of nutrients, with various nuclear receptors serving as key regulators. It has been suggested that the small intestine may also contribute to the development of IR. In normal physiological conditions, equilibrium exists between GLP-1 and GIP activity, which is continuously regulated through homeostatic mechanisms. When this balance is disrupted, abnormal GLP-1 and GIP signaling can result in persistently elevated blood glucose levels, potentially leading to the onset of IR.^122^

Investigating the mechanisms implicit in the small intestine-IR represents a analytical step towards effectively managing metabolic disorders and improving overall health outcomes. A comprehensive understanding of the molecular pathways driving small intestine-IR will not only advance our knowledge of metabolic regulation but also offer new avenues for innovations in the field of metabolic disorders.

### Large intestine in situ diabetes mellitus

Research indicates that the large intestine’s complex microbial ecosystem, known as the gut microbiome, is fundamental in regulating systemic metabolism and contributing to IR development. Changes in the composition and function of gut bacteria have been linked to increased intestinal permeability, inflammation, and changes in short-chain fatty acid production, all of which can lead to IR. Furthermore, insulin signaling in the colon plays an important role in maintaining glucose balance and overall metabolic health. Disruption of this signaling pathway can result in reduced glucose uptake and utilization by colonic epithelial cells, further contributing to systemic IR.^123^

Comparative analyses of gut microbiota have identified distinct compositional profiles between lean and obese individuals. Elevated circulating levels of lipopolysaccharide, a bacterial endotoxin originating from the intestinal lumen, are consistently observed in obesity. These findings suggest that dysbiosis, or alterations in the gut microbiota, can induce metabolic perturbations through a mechanism involving increased intestinal lipopolysaccharide translocation.^124^

Investigations into large intestinal-IR have revealed a complex interplay between the gut and metabolic conditions. By delving into the molecular pathways underlying large intestinal-IR, we can identify novel therapeutic targets aimed at enhancing colonic insulin sensitivity. This approach not only deepens our understanding of metabolic dysfunction but also paves the way for personalized therapies that account for individual gut microbiome variations. Consequently, focusing on large intestinal-IR presents a promising avenue in metabolic research, with the potential to develop strategies for addressing and managing metabolic disorders.

### Muscle in situ diabetes mellitus

In the 1960s, Randle et al.^125^ proposed one of the earliest theories to explain muscle-IR mechanisms. They determined that a sudden rise in muscle fatty acid oxidation results in citrate buildup, which hinders phosphofructokinase, a crucial glycolysis enzyme. Recent theories suggest that exposure of skeletal muscle to lipids is a primary factor in muscle-IR. The disruption of mitochondrial and endoplasmic reticulum interactions is believed to play a role in the development of muscle insulin resistance. These findings collectively indicate a significant mechanism connecting excessive muscle mitochondrial turnover to reduced insulin-stimulated glucose uptake.^126^

In skeletal muscle, IRS1 and IRS2 receptor phosphorylation regulates GLUT4 movement to the cell membrane, while Akt determines glycogen production and fat synthesis. Obese individuals experience decreased glucose uptake due to reduced IRS1 tyrosine phosphorylation and diminished Akt activation. A primary cause of lower IRS1 phosphorylation is the activation of serine kinases, which alters insulin receptor conformation. Consequently, the inability of GLUT4 to relocate in response to insulin in obese individuals leads to IR.^127^

IR manifests in muscle tissue through decreased insulin-stimulated glucose uptake, resulting from disrupted insulin signaling pathways. Research has demonstrated that muscle IR is associated with oxidative stress occurring in mitochondria.^128^

In situations of endoplasmic reticulum stress, the activation of IκB kinase (IKK) and c-Jun N-terminal kinase (JNK) is very important, as it results in the serine phosphorylation of IRS1. Additionally, the activation of Toll-like receptors (TOLL), particularly in macrophages, contributes to heightened signaling in individuals with obesity. An investigation involving animals with a mutation in TOLL-4 revealed that a high-fat diet does not induce IR, as it fails to activate IKK and JNK, thereby not stimulating insulin receptors.^129^

Furthermore, recent studies focusing on individuals who have undergone bariatric surgery have observed a significant decrease in serum levels of branched-chain amino acids, specifically valine, isoleucine, and leucine. These amino acids are known to activate the mammalian target of rapamycin, which can induce IR through the phosphorylation of IRS1 at serine residues.^130^

### Adipose Tissue in situ diabetes mellitus

Insulin serves as a critical regulator of multiple adipocyte functions. Pioneering studies in the 1990’s first established the link between inflammation and adipose tissue IR.^132131^ Macrophages play a pivotal role in amplifying inflammation within the adipose microenvironment. Once activated, their secreted cytokines exert both local and systemic effects, exacerbating inflammation and IR. The adipose tissue IR index, calculated as the product of fasting plasma insulin and free fatty acid concentrations, serves as a widely-used surrogate marker of adipose tissue IR in humans.^53^

Dysfunctional adipose tissue plays a significant role in the development of IR. In individuals with obesity, adipocytes expand in size, leading to their dysfunction. This process is accompanied by the recruitment of macrophages, which contribute to a pro-inflammatory environment through the secretion of pro-inflammatory cytokines. Additionally, fat accumulation in various organs results in lipotoxicity and disrupts the normal functioning of mitochondria, lysosomes, and the endoplasmic reticulum. These changes ultimately lead to systemic inflammation and disturbances in glucose homeostasis.^132^ Visceral adipose tissue and subcutaneous adipose tissue are both linked to IR. Consequently, the impaired adipocyte secretes an excess of free fatty acids, reactive oxygen species, and pro-inflammatory cytokines, which contribute to the development of IR through inflammation caused by lysosomal dysfunction and endoplasmic reticulum stress.

Brown adipose tissue and white adipose tissue participate in energy homeostasis. White adipose tissue is deposited under the skin and around organs, serving as a reservoir for storage of lipids and secreting adiponectin and leptin. Brown adipose tissue, on the other hand, functions in the modulation of energy balance and energy expenditure from excess food intake or in the body’s response to low temperatures. Brown adipose tissue activates thermogenesis through endocrine factors as well as due to the “browning” of white adipose tissue. The buildup of surplus white adipose tissue has negative implications for metabolic health. The stimulation of brown adipose tissue, which serves as the main organ for thermogenesis, offers advantageous effects on body fat, insulin sensitivity, and elevated lipid levels. Therefore, promoting the formation of beige adipocytes within white adipose tissue may alleviate the harmful effects associated with excess white adipose tissue and contribute to enhanced metabolic health.^133^

An additional significant point to consider is that interleukin-6 is produced by both brown and white adipocytes and plays a role in the onset of IR in individuals with obesity. Nevertheless, research has demonstrated that the transplantation of brown adipocytes into the abdominal cavity of obese exhibiting IR can fully reverse this condition.^134^ This effect is attributed to the presence of uncoupling protein 1 in brown adipocytes, which is a crucial target in the battle against diabetes and the reduction of body fat mass.

### Vascular Tissue in situ diabetes mellitus

IR impacts both arteries and arterioles affecting the endothelial and smooth muscle layers. This vascular-IR is significant in the development of vascular and related diseases. However, the exact mechanisms by which chronic inflammation induces vascular-IR remain unclear.^135^ Due to the complexity of these mechanisms, replicating the in vivo environment poses challenges. In vitro studies have been conducted to simulate acute or chronic vascular inflammatory conditions in vascular endothelial cells, and investigations have focused on vascular endothelial cells exposed to varying concentrations of TNF-α.^136^

Vascular-IR is recognized for its role in facilitating atherogenesis and the progression of advanced plaques, which contribute to cardiovascular and cerebrovascular diseases.^137^ Additionally, Vascular-IR is implicated in the development of diabetes by hindering the recruitment of capillaries and the delivery of insulin in skeletal muscle, as well as affecting glucose-induced insulin secretion from β-cells.^138^ Moreover, brain-IR, likely resulting from vascular insulin resistance, is thought to be a contributing factor to diabetic complications.

Insulin receptors are found in the endothelial cells of blood vessels, and dysfunction of these vascular endothelial cells can lead to various vascular diseases, notably atherosclerosis. The activation of nitric oxide synthase in endothelial cells, stimulated by insulin signaling, results in the production of nitric oxide, which is crucial for maintaining the integrity of the vascular barrier. This preservation is essential to prevent the accumulation of atherogenic lipoproteins within the subendothelial space. Furthermore, nitric oxide plays a significant role in mitigating the onset and progression of atherosclerosis by inhibiting the buildup of macrophages derived from monocytes in the vessel intima.^139^ Consequently, vascular-IR is a key factor in the development of vascular pathologies.

### Ovarian in situ diabetes mellitus

IR is a prevalent characteristic of polycystic ovary syndrome (PCOS), and ongoing reproductive issues indicate that IR within the ovaries may contribute to this vulnerability. The long-term consequences of IR in individuals with PCOS encompass metabolic irregularities, a heightened risk of T2DM, hypertension, cardiovascular diseases, and endometrial cancer. In polycystic ovaries, the insulin-induced stimulation of androgens in thecal cells remains intact and is even enhanced through the direct activation of insulin receptors, while the insulin-mediated uptake of glucose in granulosa cells is significantly compromised.^60^

PCOS is a prevalent condition affecting women of reproductive age, marked by persistent anovulation, elevated androgen levels, and often linked to hyperinsulinemia. Various proteins play a role in the pathophysiology of PCOS and IR, including adiponectin, apelin, vaspin, visfatin, copeptin, plasminogen activator inhibitor-1, irisin, and zonulin. Additionally, other proteins such as resistin, ghrelin, and leptin have been suggested as potential markers of IR in PCOS; however, their involvement remains a subject of debate.^140^

Insulin receptors are present on granulosa cells, theca cells, and within the ovarian stroma. Research has indicated that the binding of insulin to these receptors results in their phosphorylation, which subsequently enhances the synthesis of androgens, estrogen, and progesterone. This underscores the critical role of insulin in ovarian physiology, particularly in the context of steroidogenesis. Furthermore, hyperinsulinemia may intensify the effects of insulin-like growth factor-1 (IGF-1) by influencing IGF-1 receptors in the ovaries and modulating the production of insulin-like growth factor-binding protein-1.^141^ An additional hypothesis regarding polycystic PCOS suggests that the hyperstimulation of the ovaries, leading to elevated androgen levels, may be attributed to IR stemming from hyperinsulinemia.^142^

Thus, ovarian-IR is a significant factor in metabolic disorders such as PCOS. Although the exact mechanisms that contribute to ovarian-IR are not fully understood, various signaling pathways have been identified as relevant. Commonly observed disruptions in the IRS and PI3K pathways result in reduced glucose uptake and changes in steroid hormone production. Moreover, chronic inflammation, marked by elevated levels of TNF-α and interleukin-6, plays a role in insulin resistance by disrupting insulin signaling and enhancing serine phosphorylation of IRS proteins. Additionally, changes in the endoplasmic reticulum stress response and oxidative stress have been found to worsen IR within the ovarian context.

### Testicular in situ diabetes mellitus

Testosterone plays a significant role in the formation of both muscular and visceral adipose tissue, affecting the differentiation of pluripotent stem cells and suppressing the maturation of preadipocytes. Additionally, testosterone exerts a protective influence on pancreatic β cells, likely through mechanisms involving androgen receptors and the modulation of inflammatory cytokines. A deficiency in testosterone is linked to a higher occurrence of metabolic syndrome components, particularly the accumulation of visceral adipose tissue and the development of insulin resistance.^143^

Research indicates that insulin signaling is vital for the proper functioning of the testes, and that IR may negatively impact male reproductive health. In a healthy physiological state, insulin facilitates sperm production and maturation by promoting the synthesis of androgen-binding protein, a critical transport protein necessary for spermatogenesis. Additionally, insulin boosts the activity of Leydig cells responsible for testosterone production and encourages glucose uptake by germ cells, thereby supplying the energy required for spermatogenesis.^144^

The IR can adversely affect testicular function by interfering with insulin signaling pathways within the testes. The IR in this organ can result in diminished sperm production and maturation, along with lowered serum testosterone levels. Furthermore, IR may induce oxidative stress, which can further compromise sperm quality and fertilization potential.^145^

Testosterone treatment markedly elevated the levels of hormones associated with the hypothalamic-pituitary-gonadal axis, such as gonadotropin-releasing hormone, follicle-stimulating hormone, luteinizing hormone, testosterone itself, and insulin-like growth factor.^146^ Consequently, testicular-IR may adversely affect male reproductive health by disrupting testicular function and playing a role in the onset of metabolic syndrome.

This scoping review has some limitations that should be acknowledged. Firstly, the relatively small number of studies focusing specifically on organ-specific IR compared to the vast literature on systemic IR indicates a need for further research in this area. Secondly, the heterogeneity of study designs and methodologies used in the included studies makes it challenging to draw definitive conclusions about the prevalence and clinical significance of organ-specific IR across different populations. Finally, the focus on English-language studies may have excluded relevant research conducted in other languages. Despite these limitations, the findings of this scoping review provide a valuable foundation for future research on organ-specific IR, and the concept of “in situ diabetes mellitus” or “T6DM” offers a promising framework for understanding the heterogeneity of diabetes and guiding future investigations.

Finally, there are 17 tissues that can induce insulin resistance, and there are also 17 mechanisms that can be activated to trigger insulin resistance. The consequences of living with insulin resistance can lead to cognitive alterations, endocrine system disorders, fatty liver and pancreatic disease, cardiovascular system dysfunctions, alterations in homeostasis and coagulation, cancer, among others. Thus, once insulin resistance is established, a subclinical inflammatory process occurs, characterized by hyperinsulinemia, activation of signaling pathways, and endoplasmic reticulum stress. Some of the consequences of insulin resistance mentioned above are a result of hyperinsulinemia or alterations in the metabolism itself or the sum of these factors.

## CONCLUSION

The findings suggest that investigating organ-specific IR in the context of T2DM is a promising avenue for future research to deepen our understanding of disease pathophysiology. Thus, this scoping review answers the following question “In Situ Resistance Insulin - Localized Type 2 Diabetes Mellitus or Type 6 Diabetes Mellitus?”, emphasizing the need for targeted investigations into localized manifestations of IR and their implications for DM management strategies.

## Data Availability

All data produced in the present work are contained in the manuscript

